# Clinical and genomic epidemiology of *mcr*-*9*-carrying carbapenem-resistant Enterobacterales isolates in Metropolitan Atlanta, 2012-2017

**DOI:** 10.1101/2021.10.13.21264308

**Authors:** Ahmed Babiker, Chris Bower, Joseph D. Lutgring, Jessica Howard-Anderson, Uzma Ansari, Gillian McAllister, Michelle Adamczyk, Erin Breaker, Sarah W. Satola, Jesse T. Jacob, Michael H. Woodworth

**Affiliations:** Division of Infectious Diseases, Department of Medicine, Emory University School of Medicine, Atlanta, GA; Department of Pathology and Laboratory Medicine, Emory University School of Medicine, Atlanta, GA; Georgia Emerging Infections Program, Decatur, GA; Atlanta VA Medical Center, Decatur, GA; Foundation for Atlanta Veterans Education and Research, Decatur, GA; Division of Healthcare Quality Promotion, Centers for Disease Control and Prevention, Atlanta, Georgia, USA; Goldbelt C6, LLC, Chesapeake, Virginia, USA

**Keywords:** healthcare epidemiology, next generation sequencing, CRE, MDR, AMR

## Abstract

Colistin is a last-resort antibiotic for multidrug-resistant gram-negative infections. Recently, the ninth allele of the mobile colistin resistance (*mcr*) gene family, designated *mcr-9*, was reported. However, its clinical and public health significance remains unclear. We queried genomes of carbapenem-resistant Enterobacterales (CRE) for *mcr-9* from a convenience sample of clinical isolates collected between 2012-2017 through the Georgia Emerging Infections Program, a population- and laboratory-based surveillance program. Isolates underwent phenotypic characterization and whole genome sequencing. Phenotypic characteristics, genomic features, and clinical outcomes of *mcr-9* positive and negative CRE cases were then compared. Among 235 sequenced CRE genomes, thirteen (6%) were found to harbor *mcr-9*, all of which were *Enterobacter cloacae* complex. The median MIC, rates of heteroresistance and inducible resistance to colistin were similar between *mcr-9* positive and negative isolates. However, rates of resistance were higher among mcr-9 positive isolates across most antibiotic classes. All cases had significant healthcare exposures. The 90-day mortality was similarly high in both *mcr-9* positive (31%) and negative (7%) CRE cases. Nucleotide identity and phylogenetic analysis did not reveal geo-temporal clustering. *mcr-9* positive isolates had a significantly higher number of median [range] AMR genes (16 [4-22] vs. 6 [2-15]; *p* <0.001) compared to *mcr-9* negative isolates. Pan genome tests confirmed a significant association of *mcr-9* detection with mobile genetic element and heavy metal resistance genes. Overall, the presence of *mcr-9* was not associated with significant changes in colistin resistance or clinical outcomes but continued genomic surveillance to monitor for emergence of AMR genes is warranted.

## Introduction

With the rise of carbapenem-resistant organisms over the past few decades (1, 2), polymyxins (colistin or polymyxin B) remain last-resort antibiotics for multidrug-resistant gram-negative infections (3). While concerns regarding efficacy and nephrotoxicity (4) have relegated polymixins to the second or third line of the antibiotic armamentarium (5), these agents remain listed as critically important antibiotics by the WHO and are widely used globally.

In 2015, a colistin-resistance gene localized on a plasmid was designated mobilized colistin resistance-1 (*mcr-1*) (6). The *mcr-1* gene encodes a transferase that adds a phosphoethanolamine residue to cell membrane lipid A, altering the binding site of colistin, and consequently leading to colistin resistance (6). Since its initial description in *Escherichia coli (6)*, multiple *mcr* alleles (*mcr*-2 to *mcr*-10.1) have been described (7). The *mcr-9* allele was reported in 2019 and is most similar to *mcr-3* among previously described *mcr* alleles (8). A recent search of publicly-available sequence databases revealed a wide global distribution of *mcr-9*-harboring isolates, across six continents and in at least 9 *Enterobacterales* species (7, 9). However, it is most commonly detected among *Enterobacter* species (9).

In the US, the initial wave of carbapenem-resistant Enterobacterales (CRE) was predominantly driven by proliferation of KPC-harboring *Klebsiella pneumoniae (1, 10)*; however, as demonstrated in recent reports, a second wave of CRE in the US seems to be driven by the rise of *Enterobacter* species (1, 11). Genomic analysis indicates this second wave is associated with a high degree of clonal diversity among isolates (12). With the increasing spread of carbapenem-resistant *Enterobacter*, the dissemination of *mcr-9* is highly probable. Despite its global distribution, the impact of *mcr-9* on colistin phenotypic susceptibility remain unclear. Moreover, its association with patient clinical outcomes or potential for outbreaks of public health concern is yet to be examined.

The Centers for Disease Control and Prevention (CDC)-funded Georgia Emerging Infections Program (GA EIP) performs active, population- and laboratory-based surveillance for CRE isolated from sterile sites or urine in metropolitan Atlanta, GA (population ∼4 million). We aimed to estimate the frequency of *mcr-9* among CRE cases within the GA EIP catchment area and to compare clinical outcomes and microbiological, genomic, and clinical characteristics of *mcr-9* positive and *mcr-9* negative cases.

## Results

### The mcr-9 allele was infrequently detected among GA EIP isolates between 2012-2017

Between 2012-2017, the GA EIP identified 1,507 incident CRE cases, 716 (47.5%) were *K. pneumoniae*; 415 (27.5%), *Escherichia coli*; 270 (17.9%), *Enterobacter cloacae* complex; 72 (4.8%), *Klebsiella aerogenes (*formerly *Enterobacter)*; and 34 (2.3%) *Klebsiella oxytoca* cases. The overall crude annual CRE incidence across GA EIP increased from 4.6 to 9.6 per 100,000 population from 2012 to 2017. Carbapenem-resistant *E. cloacae* complex incidence, in particular, increased from 0.37 to 2.3 per 100,000 population during the study period. This increase coincided with revision of the CDC case surveillance definition for CRE (13). (**Figure 1 A/B, Supplementary Table 1, Supplementary Figure 1**).

**Figure 1.**
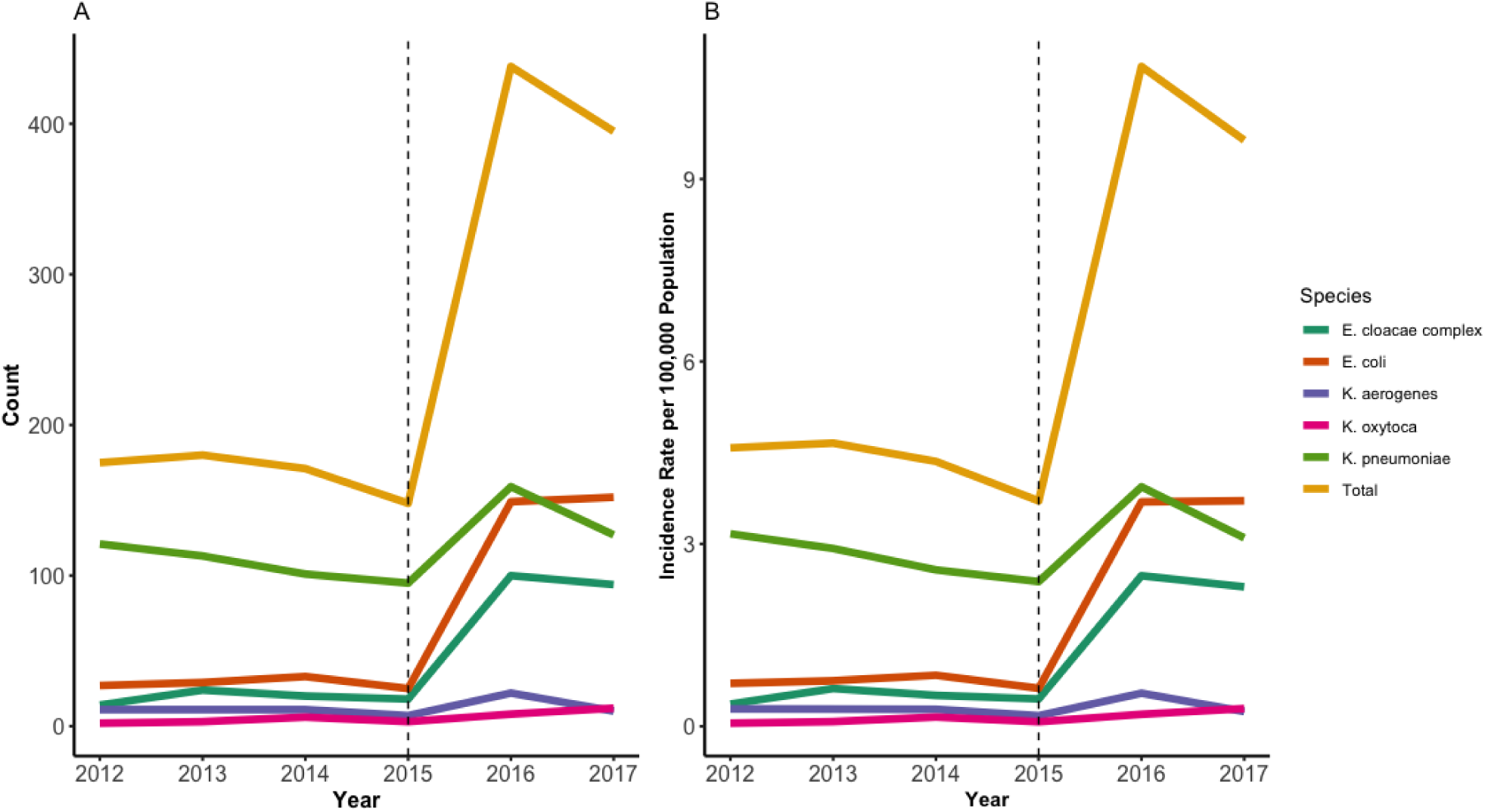
Carbapenem-resistant Enterobacterales (CRE) count (A) and crude annual incidence per 100,000 population (B) by species across from the Georgia Emerging Infections program from 2012 to 2017. Beginning in 2016, the phenotypic CRE case definition was changed to resistance to ≥1 carbapenem (now including ertapenem) with no cephalosporin parameter.

A convenience sample of 384 isolates which met the GA EIP CRE case definition was sent to the CDC for further characterization. Of the 384, 235 (61%) underwent whole genome sequencing (WGS). Among 235 sequenced CRE isolates, thirteen (6%) were found to harbor *mcr-9*, all of which were *E. cloacae* complex. All remaining sequenced *mcr-9* negative *E. cloacae* complex (n=14) were included as a comparative group, yielding a total number of 27 *E. cloacae* complex isolates.

### Microbiology Characteristics

Following collection, isolates underwent reference antimicrobial susceptibility testing by broth microdilution (BMD) at CDC. Of the *E. cloacae* complex isolates that underwent WGS from 2012-2017, twenty-two isolates (81.7%, 22/27) were confirmed to be carbapenem-resistant. Carbapenem resistance was similar among *mcr-9* positive and negative cases (84.6% (11/13) vs. 78.6% (11/14); *p*=1.00). Fluoroquinolone resistance was significantly higher among *mcr-9* positive isolates compared to *mcr-9* negative isolates (100% (13/13) vs. 57.1% (8/14); *p*=0.03). This contributed to a higher proportion of isolates being classified as difficult-to-treat resistance (DTR) (14). Overall, 48.1% (13/27) were classified as harboring DTR (14, 15). with *mcr-9* isolates having higher-rates of DTR (61.5% (8/13) vs. 35.7% (5/14) p=0.34). Similarly, rates of aminoglycoside, tetracycline, and trimethoprim-sulfamethoxazole resistance were higher among *mcr-9* positive isolates compared to *mcr-9* negative isolates (**Supplementary Table 2**).

The median [range of concentrations tested] colistin MIC for all *E. cloacae* complex isolates was 0.5 μg/mL [≤0.25->8.0]. Proportions of resistance, heteroresistance and inducible resistance were 11.1% (3/27), 48.1% (13/27) and 14.8% (4/27),respectively. There was no significant difference in colistin MIC, heteroresistance or inducible resistance between *mcr-9* positive and negative isolates (**Table 1**). Of the three *E. cloacae* complex isolates which were colistin-resistant by BMD, none were *mcr-9* positive, although one was positive for *mcr-10*.*1*.

**Table 1:** Carbapenem-resistant *E. cloacae* complex clinical and microbiological characteristics.

|  | **All (N=27)**<br>n (%) | ***mcr-9* positive**<br>**(N=13)**<br>n (%) | ***mcr-9* negative \***<br>**(N=14)**<br>n (%) | ***p* value** |
| --- | --- | --- | --- | --- |
| Culture Source |  |  |  | 0.33 |
| Urine | 24 (88.9) | 12 (92.3) | 12 (85.7) |  |
| Blood | 2 (7.4) | 1 (7.7) | 1 (7.1) |  |
| Peritoneal Fluid | 1 (3.7) | 0 (0.0) | 1 (7.1) |  |
| Year |  |  |  | 0.62 |
| 2012 | 1 (3.7) | 1 (7.7) | 0 (0.0) |  |
| 2013 | 8 (29.6) | 2 (15.4) | 6 (42.9) |  |
| 2014 | 2 (7.4) | 1 (7.7) | 1 (7.1) |  |
| 2015 | 3 (11.1) | 3 (23.1) | 0 (0.0) |  |
| 2016 | 9 (33.3) | 4 (30.8) | 5 (35.7) |  |
| 2017 | 4 (14.9) | 2 (15.4) | 2 (14.3) |  |
| Infection Onset |  |  |  | 0.71 |
| Hospital Onset | 3 (11.1) | 2 (15.4) | 1 (7.1) |  |
| Healthcare Associated-Community Onset | 10 (37.0) | 4 (30.8) | 6 (42.9) |  |
| Long Term Care Facility Onset | 14 (51.9) | 7 (53.8) | 7 (50.0) |  |
| **Microbiology Characteristics** |  |  |  |  |
| Colistin MIC (median[range])** | 0.5 [<0.25 ->0.8] | 0.5 [<0.25 -1.00] | 0.5 [<0.25 ->8.0] | 0.11 |
| Resistant | 3 (11.1) | 0 | 3 (21.4) | 0.25 |
| Heteroresistant | 13 (48.1) | 5 (38.5) | 8 (57.1) | 0.56 |
| Inducible resistance | 4 (14.8) | 2 (15.4) | 2 (14.3) | 1.00 |
| Difficult-to-Treat Resistance | 13 (48.1) | 8 (61.5) | 5 (35.7) | 0.34 |
| **Outcomes** |  |  |  |  |
| Hospitalization within 29 days after culture | 10 (37.0) | 5 (38.5) | 5 (35.7) | 0.86 |
| ICU admission ***^ | 4 (40.0) | 2 (40.0) | 2 (40.0) | 1.00 |
| In-hospital mortality ^ | 1 (10.0) | 1 (20.0) | 0 (0.0) | 0.59 |
| 90-day Mortality | 5 (18.5) | 4 (30.8) | 1 (7.1) | 0.28 |

\*1 *mcr-10* positive isolate
\*\* MIC units : µg/mL
\*\*\* Any ICU admission 7 days prior or 6 days after specimen collection
^ among 10 hospitalized patients, 5 in each group (*mcr-9* positive and negative)

### Clinical characteristics of mcr-9 positive and negative CRE cases

*E. cloacae* complex isolates were commonly isolated from urine (88.9%, 24/27), followed by blood (7.4%, 2/27) and peritoneal fluid (3.7%, 1/27). All cases had significant healthcare exposures, with 14 cases (51.9%) long term care facility onset, 10 (37.0%) healthcare-associated community onset, and 3 (11.1%) hospital onset. Ten patients (37.0%) were hospitalized at time of culture or within 29 days of CRE culture. Among the 10 hospitalized patients, 3 (30.0%) patients were admitted to the ICU within seven days of culture and one patient died during the period of hospitalization (10.0%).

Among hospitalized patients with available follow up data (n=7), 42.8% (3/7) were readmitted within 30 days. Overall unadjusted all-cause 90-day mortality was 18.5% (5/27). No clinical characteristics and outcomes were significantly different, and most were numerically similar among *mcr-9* positive and negative cases. 90-day mortality was higher among *mcr-9* positive cases (30.7% (4/13) vs. 7.1% (1/14)) compared to *mcr-9* negative cases but this was not statistically significant (*p*=0.28).

### Genomic Analysis

To expand the analytic genome set, a comparator cohort of nine publicly-available clinical and environmental carbapenem-resistant *E. cloacae* complex (three *mcr-9* positive, six *mcr-9* negative) genomes downloaded from National Center for Biotechnology Information (NCBI). Review of associated metadata reveled isolates to have been collected between January 2012 and December 2016 at the National Institutes of Health (NIH) clinical center (Bethesda, MD) (PRJNA430442) (16). These genomes were combined with the 27 GA EIP isolates, resulting in 36 isolate draft genomes included in further comparative genomic analyses.

The 13 *mcr-9* positive *E. cloacae* complex isolates were obtained from 10 distinct GA EIP facilities across the study period (2012-2017). Average nucleotide identity pairwise comparisons revealed three distinct clusters (**Figure 2**). This clustering did appear to be related to geographical location or year of isolation. NIH isolates clustered with GA EIP isolates, and GA EIP isolates did not cluster by facility or year (**Supplementary Figure 2**). Their distribution throughout the genome phylogeny suggested that the *mcr-9* positive isolates were largely genetically distinct from one another.

**Figure 2.**
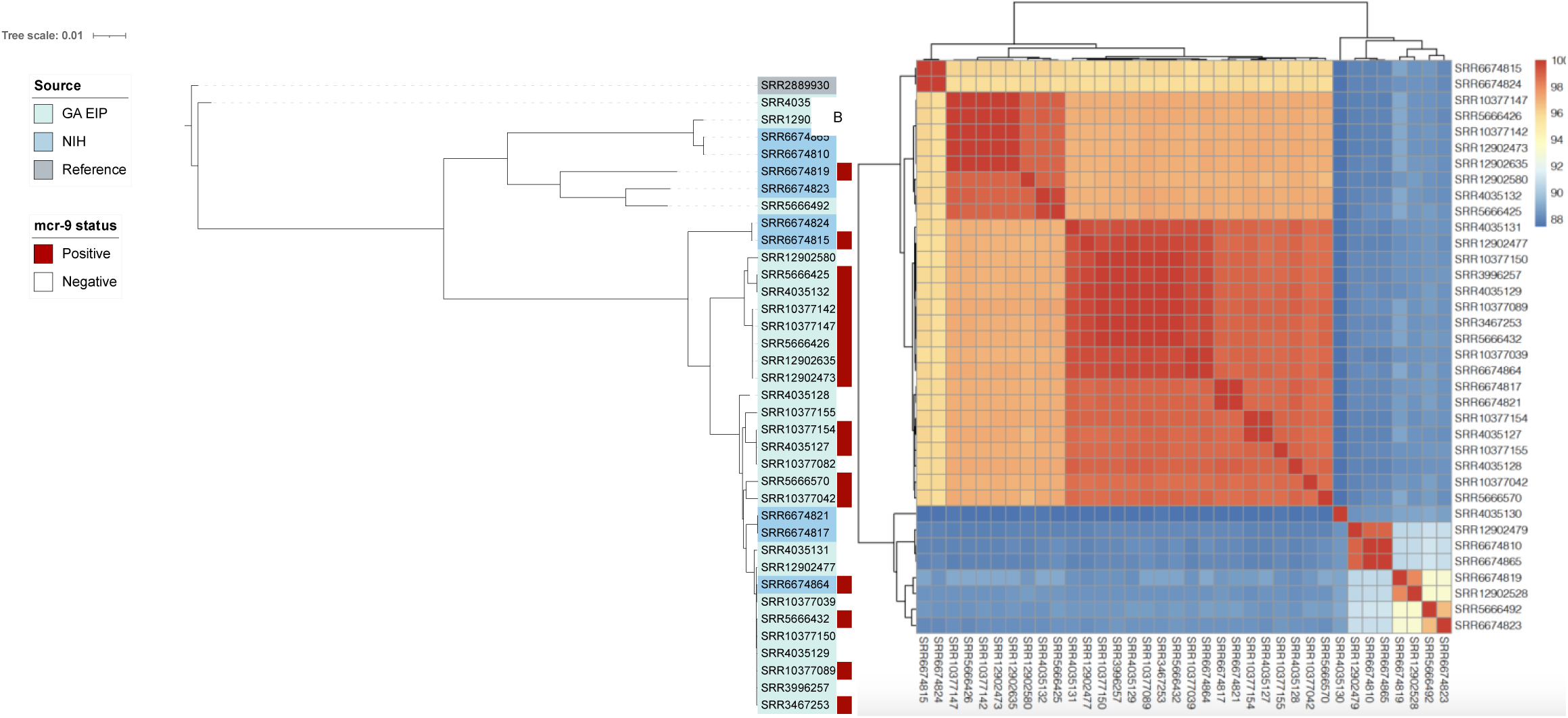
Phylogeny (A) and average nucleotide identity heatmap (B) of *mcr-9* positive (n=13) and *mcr-9* negative (n=14) *E. cloacae* complex genomes from Georgia Emerging Infection program in addition to 9 available *E. cloacae* complex genomes (three *mcr-9* positive, six *mcr-9* negative) from the National Institutes of Health. A phylogenetic tree based on a core gene alignment containing 1,904 genes defined using Roary v3.13.0. was generated using IQtree v2.0.3. A maximum likelihood tree was generated by running 1,000 bootstrap replicates under the generalized time-reversible model of evolution. The tree was visualized and annotated using Interactive Tree of Life (iTOL) v4. Pairwise comparisons of average nucleotide identity on the assembled genomes were performed with the Mashmap method using fastANI v1.32. Abbreviations: GA EIP: Georgia Emerging Infections Program, NIH: National Institutes of Health,

Median [range] AMR gene content (excluding *mcr-9*) was significantly higher among *mcr-9* positive isolates compared to *mcr-9* negative isolates (16 [4-22] vs. 6 [2-15]; p <0.001) (**Figure 3**). Among the three isolates with elevated colistin MICs, no point mutations in *pmrA/pmrB*, a two-component system regulator of lipopolysaccharide (the target site of colistin) modifications, were detected. Pan-genome-wide association tests revealed a significant association of *mcr-9* detection with the detection of mobile genetic element (MGE)-associated genes such as *repB, parM, hns2;* heavy metal resistance (HMR) genes such as *arsC2, arsB2, fieF2, pcoE2* and *merA*; and virulence genes such *hipA* (**Table 2, Supplementary Table 2, Supplementary Figure 3**). Taken together, these comparative genomic analyses across two sites with *mcr-9* positive *E. cloacae* complex isolate draft genomes confirmed the colocalization of *mcr-9* with plasmid-mobilized heavy metal resistance genes but did not provide evidence of a high-identity outbreak cluster in space or time.

**Figure 3.**
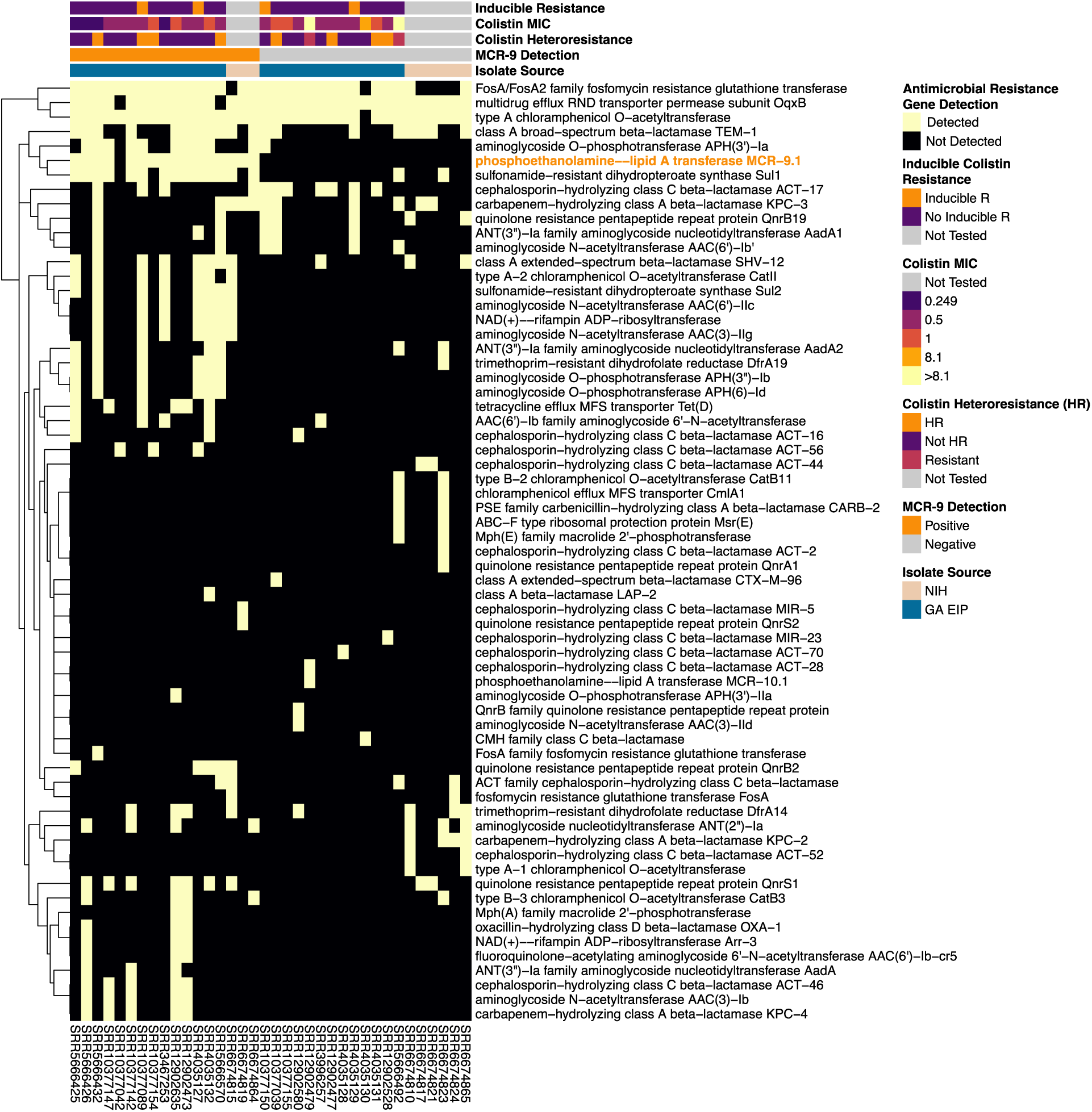
Antimicrobial resistance gene heatmap of *mcr-9* positive (n=13) and *mcr-9* negative (n=14) *E. cloacae* complex genomes from Georgia Emerging Infections program in addition to 9 available *E. cloacae* complex genomes (three *mcr-9* positive, six *mcr-9* negative) from the National Institutes of Health. Genomes were annotated using Prodigal v2.6.3 and antimicrobial resistance gene content was assessed using AMRFinder Antimicrobial resistance gene presence/absence heatmaps were created using the package pheatmap on R version 4.0.2 (Vienna, Austria) and the RStudio interface version 1.3.1073 (Boston, MA, USA).

**Table 2.** Highest ranking genes for association with *mcr-9* presence.

| Gene* | Comment | Odds Ratio | Bonferroni<br>adjusted *p*-<br>value |
| --- | --- | --- | --- |
| *smc* | Chromosome partition protein Smc | ∞ | 1.60E-06 |
| *dcm2* | DNA-cytosine methyltransferase | ∞ | 1.60E-06 |
| *pcoE2* | putative copper-binding protein PcoE | ∞ | 2.88E-05 |
| *hns2* | DNA-binding protein H-NS, plasmid | ∞ | 3.36E-05 |
| group_10390 | Tn3 family transposase ISEc63 | ∞ | 3.36E-05 |
| *hipA* | Serine/threonine-protein kinase toxin HipA | ∞ | 3.36E-05 |
| *rcnR2* | Transcriptional repressor RcnR | ∞ | 0.00030219 |
| *hha2* | Hemolysin expression-modulating protein Hha | ∞ | 0.00036934 |
| *uvrD2* | DNA helicase II | ∞ | 0.00036934 |
| parM | Plasmid segregation protein | ∞ | 0.00036934 |
| *higB-1* | Toxin HigB-1 | ∞ | 0.00036934 |
| group_1846 | Tn3 family transposase ISEc63 | ∞ | 0.00036934 |
| *dam2* | DNA adenine methylase | ∞ | 0.00036934 |
| *repB* | RepFIB replication protein A | ∞ | 0.00036934 |
| *traC* | Protein TraC | ∞ | 0.00036934 |
| group_7173 | Stable plasmid inheritance protein | ∞ | 0.00036934 |
| *yjcD* | Putative ATP-dependent DNA helicase YjcD | 304 | 0.00054522 |
| *umuD2* | Protein UmuD | ∞ | 0.00283164 |
| *dsbC_2* | Thiol:disulfide interchange protein DsbC | ∞ | 0.00283164 |
| *virB* | Virulence regulon transcriptional activator VirB | ∞ | 0.00283164 |
| *umuC_3* | Protein UmuC | ∞ | 0.00283164 |
| group_7174 | IS110 family transposase ISEsa2 | 142.5 | 0.00540586 |
| *merA* | Mercuric reductase | ∞ | 0.01698985 |
| *fieF_2* | Ferrous-iron efflux pump FieF | 90.7 | 0.03283809 |
| *arsB_2* | Arsenical pump membrane protein | 67.5 | 0.04368339 |
| group_7063 | ISNCY family transposase ISEsa1 | 67.5 | 0.04368339 |
| group_8953 | ISNCY family transposase ISBcen27 | 67.5 | 0.04368339 |
| *arsH* | NADPH-dependent FMN reductase ArsH | 67.5 | 0.04368339 |
| *arsC2* | Arsenate reductase | 67.5 | 0.04368339 |

hypothetical proteins not included

## Discussion

Among 235 CRE isolates collected through a comprehensive, population-based surveillance program targeting the most common CRE species, we found a low prevalence of *mcr-9*, all of which were detected in *Enterobacter cloacae* isolates. Our phylogenetic analyses revealed a genetically diverse *mcr-9* positive CRE population, suggesting sporadic carriage rather than clonal spread. Using multi-modal phenotypic testing, we were unable to detect impacts of *mcr-9* on colistin susceptibility, however genomic analysis revealed an association with increased AMR, HMR and virulence genes. In addition, our *mcr-9*-containing CRE isolates were exclusively acquired in healthcare settings, with a trend towards increased mortality. Since their initial description, recognition of *mcr* genes associated with colistin resistance has spread rapidly across the globe (7). Our study of *mcr-9*-harboring CRE cases provides unique insights into the phenotypic and genomic implications of *mcr-9*, and is one of the first to examine clinical outcomes.

Whether *mcr-9* confers colistin resistance has been debated (17). The first isolate identified to harbor *mcr-9* was also susceptible to colistin but the allele was found to confer resistance to colistin when cloned into a colistin-susceptible *E. coli* and expressed under the control of an IPTG-induced promoter. However, this was only at 1, 2, and 2.5 mg/L, but not at 5 mg/L of colistin (8). Kieffer *et al*. later reported that *mcr-9* expression was inducible in the presence of colistin when located upstream of the two-component sensor kinase system *qseBC (17)*. This two-component signaling network allows bacteria to sense and respond to their changing environments. In particular, the *qseC* and *qseB* genes encode a histidine kinase sensor (*qseC*) and its cognate partner (*qseB*). The *qseBC* system has been shown to interact with *pmrA/pmrB*, to induce resistance to the colistin(18). However, a study of *mcr-9*-containing isolates from retail meat conducted by the National Antimicrobial Resistance Monitoring System (NARMS), found all 105 isolates (99 *S. enterica* and 6 *E. coli*) tested to be susceptible to colistin, including 10 isolates with *qseBC (19)*, indicating that the previously demonstrated impact of *qseBC* on *mcr-9* expression and colistin resistance may be dependent on strain backgrounds, as originally demonstrated in *E. coli (17)*. Among clinical CRE isolates, we found the presence of *mcr-9* was not associated with frank or inducible colistin resistance. Furthermore, our study is the first to examine the association of *mcr-9* with heteroresistance. Heteroresistance is a largely unrecognized form of antibiotic resistance where only a subset of cells within a bacterial population are resistant to a given drug (20). These resistant cells can be selected for in the presence of the antibiotic and cause colistin treatment failures *in-vivo* (21). In a multisite surveillance study of colistin heteroresistance among CRE, *Enterobacter spp*. and in particular *E. cloacae*, displayed the highest proportion of colistin heteroresistance (22). However, here we found no association between *mcr-9* and colistin heteroresistance.

Carbapenem-resistant gram-negatives are a public health threat broadly prioritized by public health organizations (23). Given the limited therapeutic options, morbidity and mortality rates are increased disproportionately when compared to infections caused by susceptible bacteria. We observed high 90-day mortality rates, but these were similar to reported CRE mortality rates at other US academic centers (1). However, there was a non-significant trend for higher mortality among *mcr-9* positive isolates. This association should be further evaluated in larger studies, with adjustment for potentially-confounding variables associated with mortality such as severity of illness, age and comorbidities as our small size may underestimate differences in mortality (1). This finding may be related to the increase in phenotypic resistance and AMR gene content associated with *mcr-9*. A similar finding was previously reported describing 1,035 *mcr-9*-containing isolates in which 97% (1003/1035) were classified as MDR (7). This increased AMR gene content, renders isolates not only carbapenem-resistant, but also with DTR, further limiting therapeutic options (14). DTR is a clinically relevant and functional classification of resistance which signifies *in vitro* resistance to all high-efficacy, low-toxicity (or first-line) agents and is associated with worse clinical outcomes when compared to carbapenem-resistant phenotypes (14). Moreover, genome-wide association studies, revealed a difference in key virulence genes such *hipA*, a eukaryote-like serine threonine kinase that inhibits cell growth and induces bacterial persistence (24).

We found the presence of *mcr-9* to be associated with HMR genes such *arsA, merA*, conferring arsenic and mercury resistance, respectively. There has been increasing evidence for the co-selection of AMR and HMR genes through either co-resistance or cross-resistance (25). Co-resistance occurs when AMR and HMR genes are carried on the same mobile genetic element. IncH12 plasmids, which frequently harbor HMR genes (8), have been found to be the predominant replicon type carrying *mcr-9* and frequently demonstrated in our isolates. Hospital wastewater is an increasingly recognized reservoir for resistant gram-negative organisms that cause healthcare-associated infections (26) and HMR genes may allow for continued persistence in the environment (27). While the community setting is starting to represent an increased source of multidrug-resistant infections (28), the healthcare setting still represents a major risk for MDR acquisition, as was seen among our cohort.

Our study combines detailed epidemiological, clinical, phenotypic, and genomic data to examine the significance of *mcr-9* but has some limitations. Firstly, we could not do a full interrogation of *mcr-9*-containing plasmids, due to limitations of short read sequencing. However, prior studies have significantly characterized the genomic background of *mcr-9*-containing plasmids (9). Second, our study did not include *Salmonella* species which are a major reservoir for *mcr-9* or other *Enterobacterales* species such as *Citrobacter* (7), and our findings may not be generalizable to these species. However, our dataset of 235 includes the most common and significant clinical CRE species (1, 23) and is one of few studies carried out on clinical human isolates (19). Third, while we assessed for presence of the *pmrA/pmrB* regulatory system (29), we did not include assessment of the two-component system *qseBC* (8) or testing conditions (such as the use of cation-adjusted Mueller-Hinton broth) (30) which have been shown to influence *mcr-9* expression and the colistin MIC results, respectively. Fourth, while our overall cohort is from a population-based surveillance program, the collected and sequenced isolates represent a convenience sample which may limit generalizability and our sample size was not powered to detect clinical outcomes.

In conclusion, *mcr-9* may not have actionable public health implications as other *mcr* alleles, most of which consistently display colistin resistance. However, given the increased AMR and HMR gene content, continued genomic surveillance of multidrug-resistant organisms to monitor for the emergence of AMR genes such as *mcr-9* is prudent. Especially as changes in the up or downstream genetic context or the accumulation of mutations may impact its ability to confer colistin resistance.

## Methods

CRE cases were identified by routine queries on automated testing instruments in the clinical labs that serve residents of the GA EIP catchment area. Clinical characteristics were obtained through medical record review, all-cause mortality data was obtained through the Georgia Vital Statistics records, and hospital readmission data was obtained through the Department of Public Health’s hospital discharge datasets. Georgia EIP surveillance activities are reviewed and approved by the Emory University Institutional Review Board.

From 2012-2015, a CRE case was defined as an isolate of *E. coli, E. cloacae* complex, *K. (*formally *Enterobacter) aerogenes, K. pneumoniae*, or *K. oxytoca* collected from a normally sterile body site (e.g., bloodstream) or urine that tested non-susceptible to ≥1 carbapenem (imipenem, meropenem, or doripenem) and resistant to all 3^rd^ generation cephalosporins tested (ceftriaxone, ceftazidime, and cefotaxime) by testing performed at the local collection microbiology laboratory. Beginning in 2016, the phenotypic case definition was changed to resistance to ≥1 carbapenem (now including ertapenem) with no cephalosporin parameter. Antibiotic susceptibility interpretations were determined using the current Clinical and Laboratory Standards Institute breakpoints (31). Fluoroquinolone resistance was defined as non-susceptibility (intermediate or resistant) to ≥1 fluoroquinolone. DTR was defined as intermediate or resistant to all reported agents in carbapenem, β-lactam, and fluoroquinolone categories (14, 15).

An incident CRE case was defined as the first CRE isolate from a patient during a 30-day period that met the surveillance definition. All incident CRE cases underwent medical record review using a standardized abstraction form. Both inpatient and outpatient medical records were reviewed for patient demographics, underlying clinical comorbidities, location of culture collection, specimen source, associated infectious syndromes, relevant health care exposures and patient outcomes. 90-day mortality was determined based by matching to vital records.

A convenience sample of CRE isolates are collected annually and submitted to the CDC for further characterization. Isolates that are collected and matched to an incident case with a completed case report form are eligible for shipment. All isolates undergo repeat reference BMD at the CDC followed by whole genome sequencing using an Illumina MiSeq benchtop sequencer.

All *mcr-9* positive and a comparative control group of *mcr-9* negative *E. cloacae* complex isolates underwent additional population analysis profiling and inducible resistance testing at the Emory Investigational Clinical Microbiology Core as previously described (20). Briefly, all isolates were tested via the population analysis profile (PAP) method. This consists of plating overnight cultures of each isolate onto solid Muller-Hinton (MH) agar with or without colistin concentrations of 0.5, 1, 2, 4, 16, 32, and 100 μg/ml. Surviving colonies were enumerated and used to detect colistin-resistant subpopulations characteristic of heteroresistance (20). Inducible resistance testing was performed as previously described (32). Briefly, a single colony of each clinical isolate was grown in Muller-Hinton (MH) broth overnight at 37°C, and cultures were diluted 1:100 in MH broth containing serially increasing concentrations of colistin, starting at the one-half MIC value of the respective isolate, and doubling every 24 hours until bacterial growth was completely inhibited (with no bacteria growth after spreading 100 µL of the culture on MH agar plates supplemented with corresponding concentration of colistin). The concentration of colistin at which bacterial growth was completely inhibited was recorded as the final colistin concentration.

### Bioinformatic Methods

Fastq files of *Enterobacter cloacae* complex isolates of interest were downloaded from the Sequence Read Archive (SRA) repository maintained by the National Center for Biotechnology Information (NCBI) using the fasterq-dump tool from the SRA Toolkit V. 2.5.7 (http://ncbi.github.io/sra-tools/). Illumina reads were quality filtered using Trimmomatic (33) and assembled *de novo* using SPAdes v3.13 (34). Pairwise comparisons of average nucleotide identity on the assembled genomes were performed with the Mashmap method using fastANI v1.32 (35). Gene sequences were predicted with Prodigal v2.6.3 (36) and annotated with Prokka v1.14.6 (37). Antimicrobial resistance and virulence gene content was assessed using AMRFinder Plus (38).

Presence of plasmids and point mutations in housekeeping genes associated with colistin resistance were assessed using the ResFinder and PlasmidFinder web interface with default settings and the *E. coli* database (39, 40). AMR gene presence/absence heatmaps were created using the package pheatmap on R version 4.0.2 (Vienna, Austria) and the RStudio interface version 1.3.1073 (Boston, MA, USA). Core genes were defined using Roary v3.13 (41). A phylogenetic tree based on a core gene alignment containing 1942 genes was generated using IQtree v2.0.3 (42). A maximum likelihood tree was generated by running 1,000 bootstrap replicates under the generalized time-reversible model of evolution. The tree was visualized and annotated using Interactive Tree of Life (iTOL) v4 (43). Pan-genome wide comparison of core genomes of *mcr-9* positive to *mcr-9* negative genomes was completed using scoary (44).

### Statistical analysis

Annual incidence rates for CRE cases were calculated using the annual US census estimates of the surveillance area population as the denominator. Descriptive analyses were performed to summarize specimen information, health care exposures, outcomes, and microbiological results of incident cases; χ2 and Wilcoxon rank sum tests were used to compare groups when applicable. Gene differences were assessed by a *p*-value adjusted with Bonferroni’s method for multiple comparisons correction. Statistical analysis was performed using R version 4.0.2 (Vienna, Austria) and the Rstudio interface version 1.3.1073 (Boston, MA, USA). A 2-sided P value of <.05 was considered statistically significant.

## Data Availability

All data produced in the present study are available upon reasonable request to the authors

## Acknowledgments

We would like to thank the EIP and MuGSI staff and the CDC laboratory staff for their dedication and work to collect, validate, clean, and maintain the data used in this analysis. We would like to thank Alison Laufer Halpin for supervising the sequencing efforts of these isolates.

## Supplementary Material

**Supplemental Table 1:** Definitions of carbapenem-resistant Enterobacterales (CRE) and difficult-to-treat resistance (DTR) used during the study period.

| Definition | Period | Species | Susceptibility Phenotype |
| --- | --- | --- | --- |
| Initial GA EIP surveillance definition | 2011–2015 | *Escherichia coli*<br>*Klebsiella pneumoniae*<br>*Klebsiella oxytoca*<br>*Enterobacter cloacae* complex<br>*Klebsiella aerogenes*<sup>a</sup> | <u>Intermediate or resistant to:</u><br>- Imipenem (MIC $\geq 2$ $\mu\text{g/mL}$), or<br>- Meropenem (MIC $\geq 2$ $\mu\text{g/mL}$), or<br>- Doripenem (MIC $\geq 2$ $\mu\text{g/mL}$)<br><u>AND resistant to<sup>b</sup>:</u><br>- Ceftazidime (MIC $\geq 16$ $\mu\text{g/mL}$), and<br>- Ceftriaxone (MIC $\geq 4$ $\mu\text{g/mL}$), and<br>- Cefotaxime (MIC $\geq 4$ $\mu\text{g/mL}$) |
| Revised GA EIP surveillance definition | 2016–present | *Escherichia coli*<br>*Klebsiella pneumoniae*<br>*Klebsiella oxytoca*<br>*Enterobacter cloacae* complex<br>*Klebsiella aerogenes*<sup>a</sup> | <u>Resistant to:</u><br>- Imipenem (MIC $\geq 4$ $\mu\text{g/mL}$), or<br>- Meropenem (MIC $\geq 4$ $\mu\text{g/mL}$), or<br>- Doripenem (MIC $\geq 4$ $\mu\text{g/mL}$), or<br>- Ertapenem (MIC $\geq 2$ $\mu\text{g/mL}$) |
| DTR | 2011–2017 | *Enterobacter cloacae*<br>complex | Intermediate or resistant to all tested<br>and reported agents in carbapenem,<br>β-lactam, and fluoroquinolone<br>categories |

a. Formerly *Enterobacter aerogenes*
b. If the antibiotics were tested
Abbreviations: GA EIP: Georgia Emerging Infections Program; DTR: difficult-to-treat
resistance; MIC: minimum inhibitory concentrations

**Supplementary Table 2:**
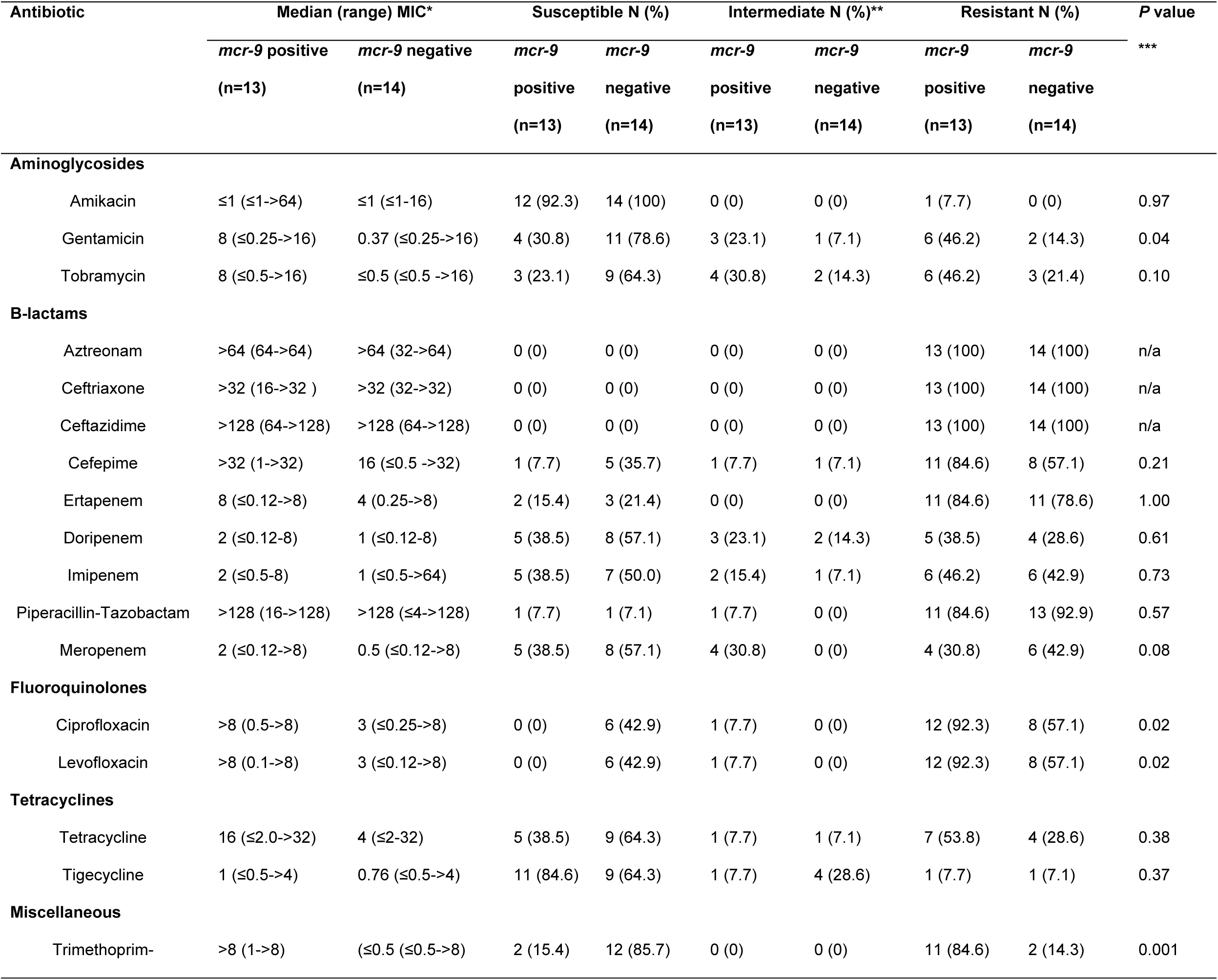
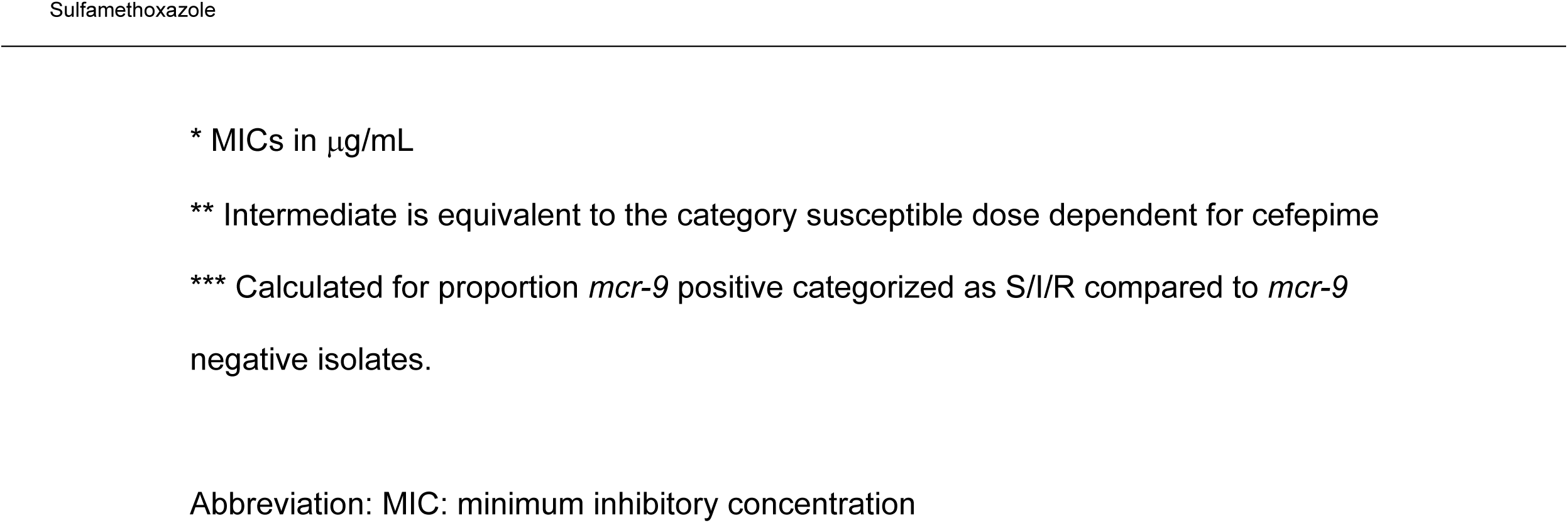
Antimicrobial Susceptibility testing of Carbapenem-resistant *E. cloacae* complex by *mcr-9* status.

| Antibiotic | Median (range) MIC* |  | Susceptible N (%) |  | Intermediate N (%)** |  | Resistant N (%) |  | P value |
| --- | --- | --- | --- | --- | --- | --- | --- | --- | --- |
|  | *mcr-9* positive | *mcr-9* negative | *mcr-9* | *mcr-9* | *mcr-9* | *mcr-9* | *mcr-9* | *mcr-9* |  |
|  | (n=13) | (n=14) | positive | negative | positive | negative | positive | negative |  |
|  |  |  | (n=13) | (n=14) | (n=13) | (n=14) | (n=13) | (n=14) |  |
| Aminoglycosides |  |  |  |  |  |  |  |  |  |
| Amikacin | ≤1 (≤1->64) | ≤1 (≤1-16) | 12 (92.3) | 14 (100) | 0 (0) | 0 (0) | 1 (7.7) | 0 (0) | 0.97 |
| Gentamicin | 8 (≤0.25->16) | 0.37 (≤0.25->16) | 4 (30.8) | 11 (78.6) | 3 (23.1) | 1 (7.1) | 6 (46.2) | 2 (14.3) | 0.04 |
| Tobramycin | 8 (≤0.5->16) | ≤0.5 (≤0.5 ->16) | 3 (23.1) | 9 (64.3) | 4 (30.8) | 2 (14.3) | 6 (46.2) | 3 (21.4) | 0.10 |
| B-lactams |  |  |  |  |  |  |  |  |  |
| Aztreonam | >64 (64->64) | >64 (32->64) | 0 (0) | 0 (0) | 0 (0) | 0 (0) | 13 (100) | 14 (100) | n/a |
| Ceftriaxone | >32 (16->32 ) | >32 (32->32) | 0 (0) | 0 (0) | 0 (0) | 0 (0) | 13 (100) | 14 (100) | n/a |
| Ceftazidime | >128 (64->128) | >128 (64->128) | 0 (0) | 0 (0) | 0 (0) | 0 (0) | 13 (100) | 14 (100) | n/a |
| Cefepime | >32 (1->32) | 16 (≤0.5 ->32) | 1 (7.7) | 5 (35.7) | 1 (7.7) | 1 (7.1) | 11 (84.6) | 8 (57.1) | 0.21 |
| Ertapenem | 8 (≤0.12->8) | 4 (0.25->8) | 2 (15.4) | 3 (21.4) | 0 (0) | 0 (0) | 11 (84.6) | 11 (78.6) | 1.00 |
| Doripenem | 2 (≤0.12-8) | 1 (≤0.12-8) | 5 (38.5) | 8 (57.1) | 3 (23.1) | 2 (14.3) | 5 (38.5) | 4 (28.6) | 0.61 |
| Imipenem | 2 (≤0.5-8) | 1 (≤0.5->64) | 5 (38.5) | 7 (50.0) | 2 (15.4) | 1 (7.1) | 6 (46.2) | 6 (42.9) | 0.73 |
| Piperacillin-Tazobactam | >128 (16->128) | >128 (≤4->128) | 1 (7.7) | 1 (7.1) | 1 (7.7) | 0 (0) | 11 (84.6) | 13 (92.9) | 0.57 |
| Meropenem | 2 (≤0.12->8) | 0.5 (≤0.12->8) | 5 (38.5) | 8 (57.1) | 4 (30.8) | 0 (0) | 4 (30.8) | 6 (42.9) | 0.08 |
| Fluoroquinolones |  |  |  |  |  |  |  |  |  |
| Ciprofloxacin | >8 (0.5->8) | 3 (≤0.25->8) | 0 (0) | 6 (42.9) | 1 (7.7) | 0 (0) | 12 (92.3) | 8 (57.1) | 0.02 |
| Levofloxacin | >8 (0.1->8) | 3 (≤0.12->8) | 0 (0) | 6 (42.9) | 1 (7.7) | 0 (0) | 12 (92.3) | 8 (57.1) | 0.02 |
| Tetracyclines |  |  |  |  |  |  |  |  |  |
| Tetracycline | 16 (≤2.0->32) | 4 (≤2-32) | 5 (38.5) | 9 (64.3) | 1 (7.7) | 1 (7.1) | 7 (53.8) | 4 (28.6) | 0.38 |
| Tigecycline | 1 (≤0.5->4) | 0.76 (≤0.5->4) | 11 (84.6) | 9 (64.3) | 1 (7.7) | 4 (28.6) | 1 (7.7) | 1 (7.1) | 0.37 |
| Miscellaneous |  |  |  |  |  |  |  |  |  |
| Trimethoprim- | >8 (1->8) | (≤0.5 (≤0.5->8) | 2 (15.4) | 12 (85.7) | 0 (0) | 0 (0) | 11 (84.6) | 2 (14.3) | 0.001 |

\* MICs in µg/mL
\*\* Intermediate is equivalent to the category susceptible dose dependent for cefepime
\*\*\* Calculated for proportion *mcr-9* positive categorized as S/I/R compared to *mcr-9* negative isolates.
Abbreviation: MIC: minimum inhibitory concentration

**Supplementary Table 3:**
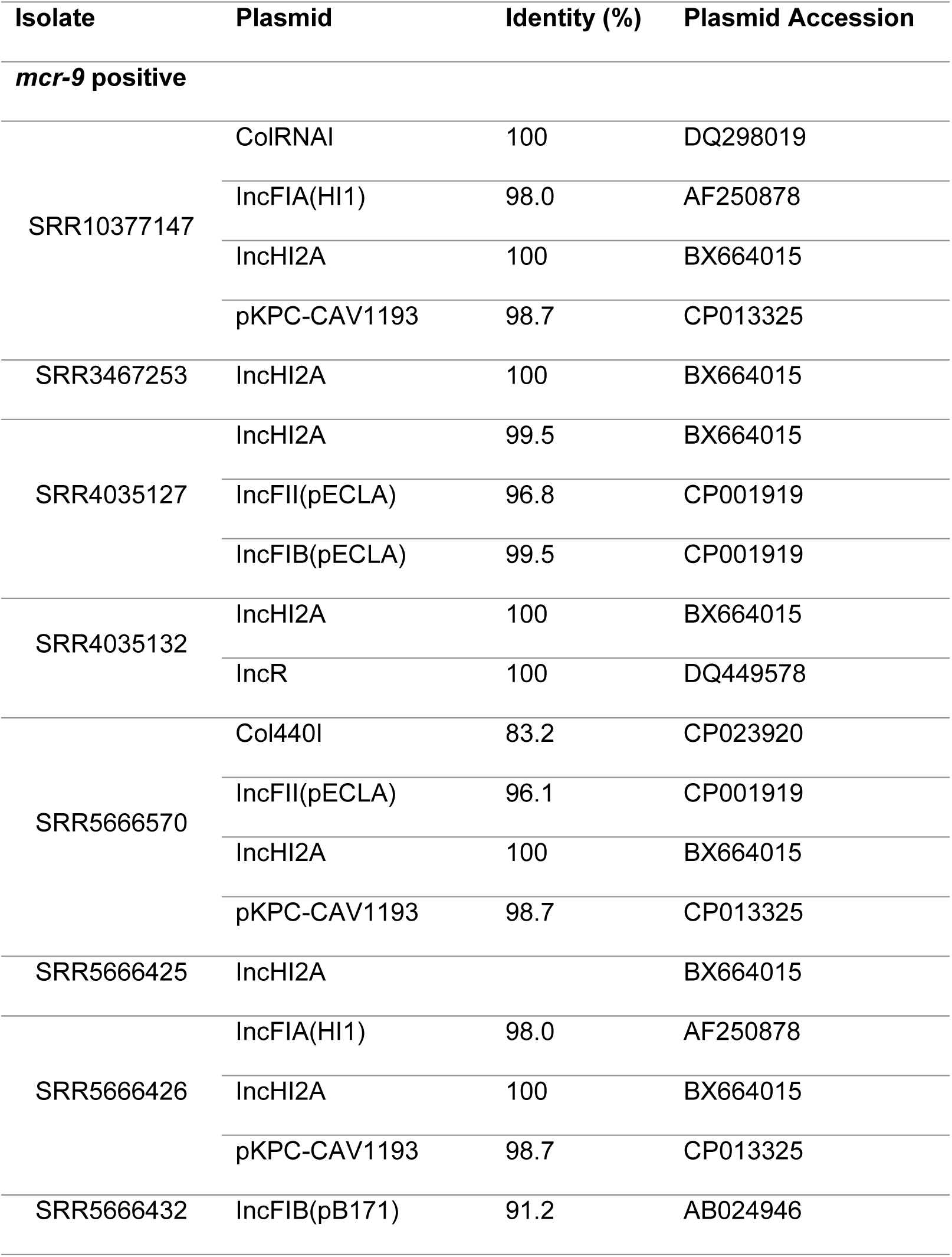
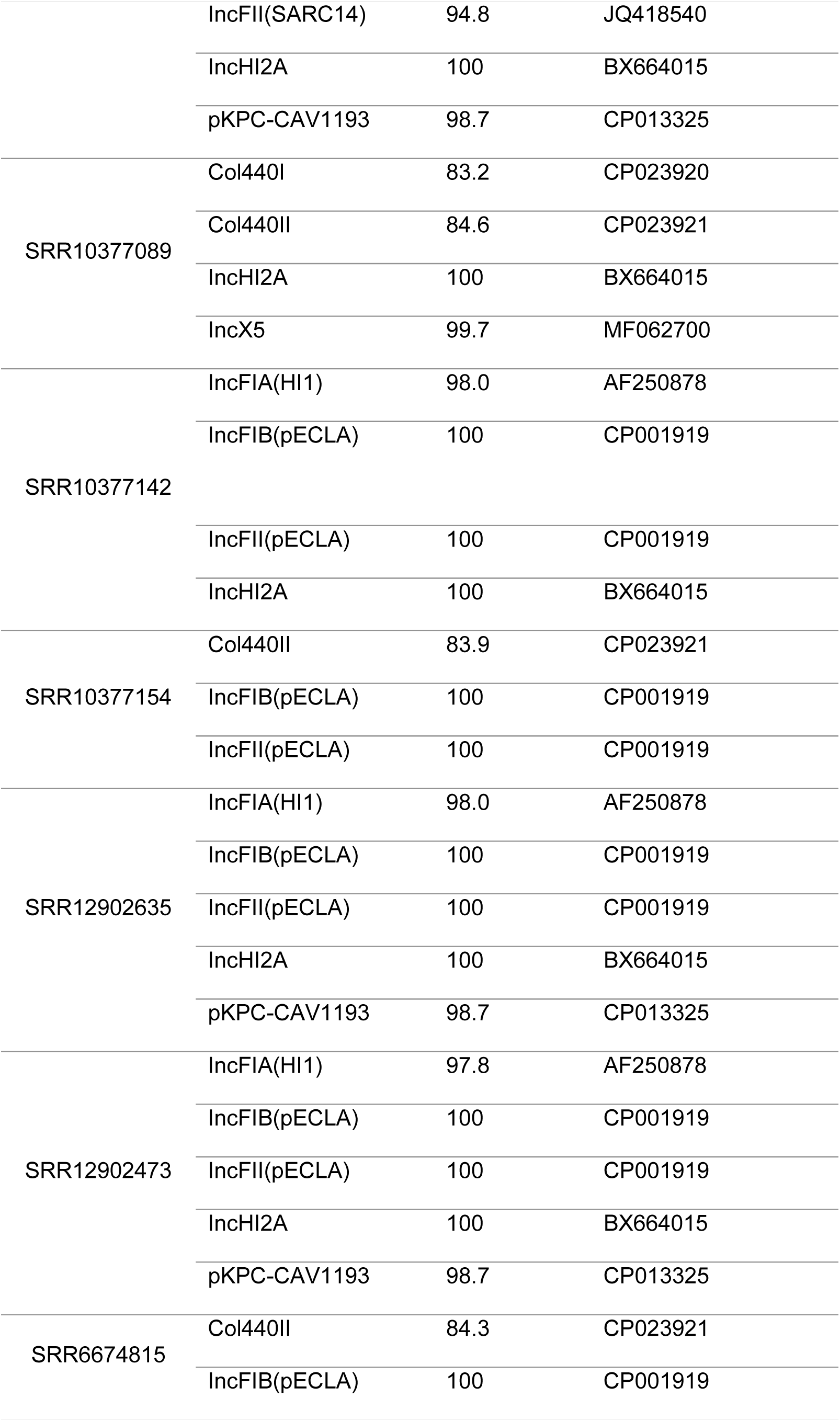
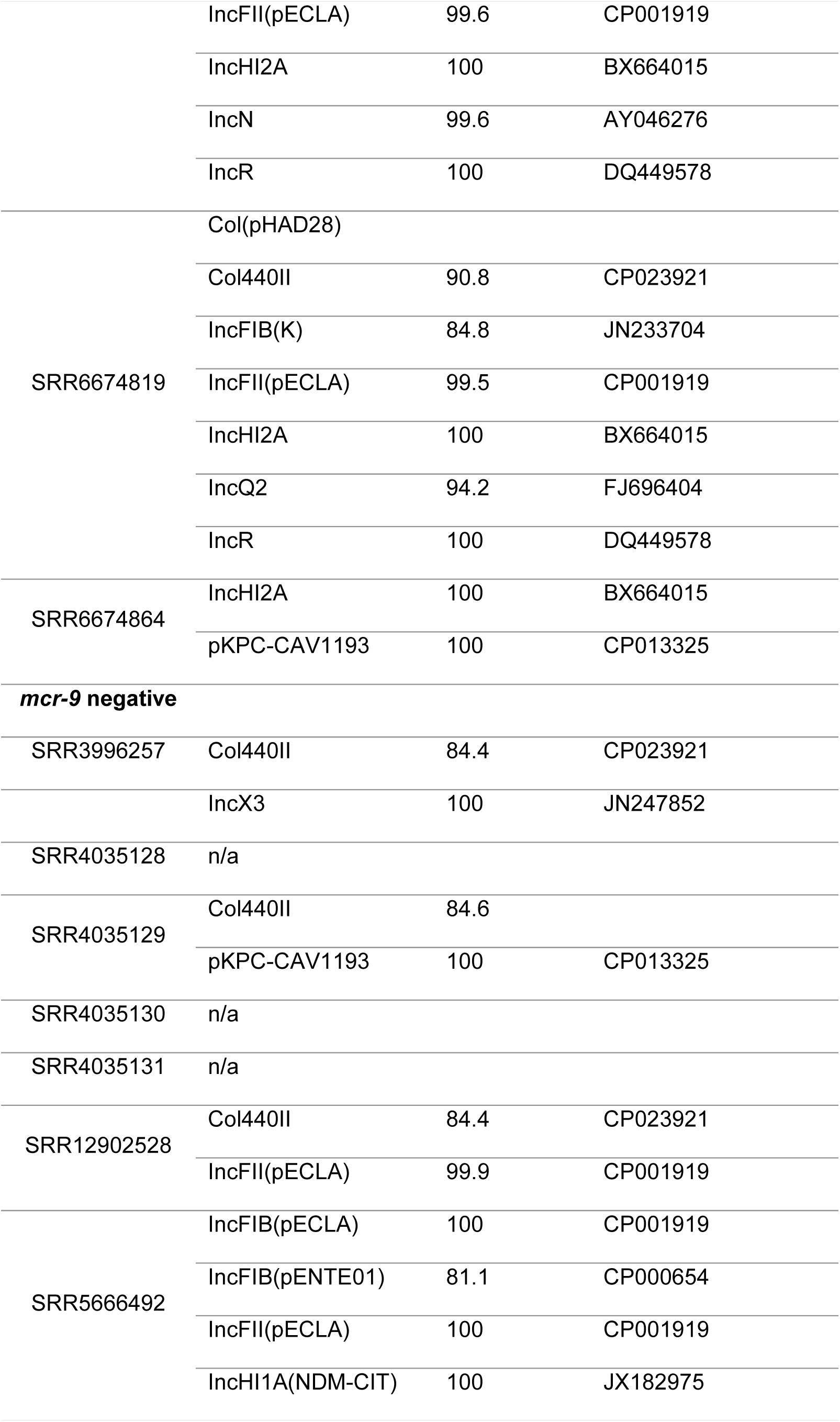
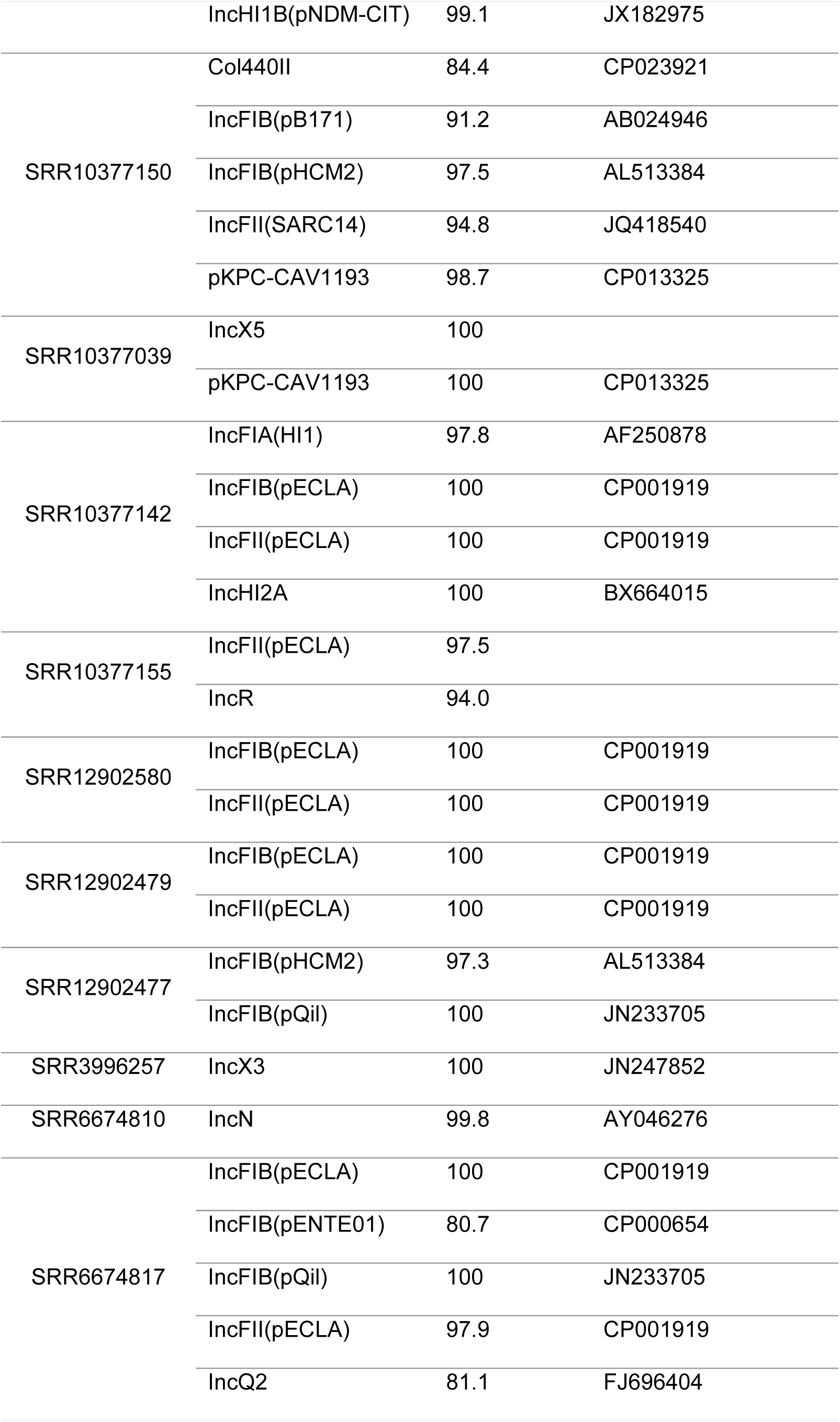
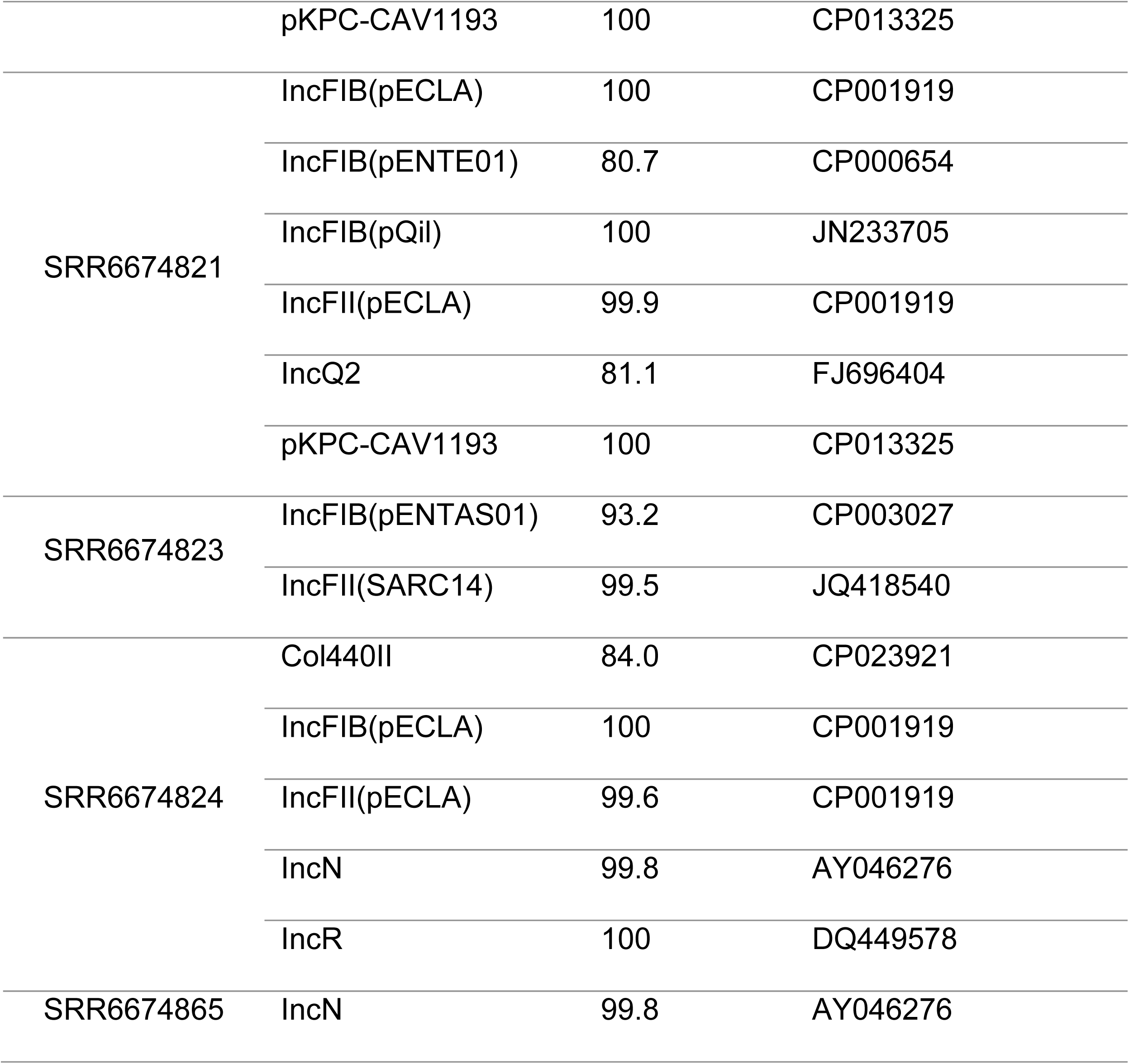
Plasmid contents of Carbapenem-resistant *E. cloacae* complex genomes *by mcr-9 status*.

## Supplementary Figures

**Supplementary Figure 1.**
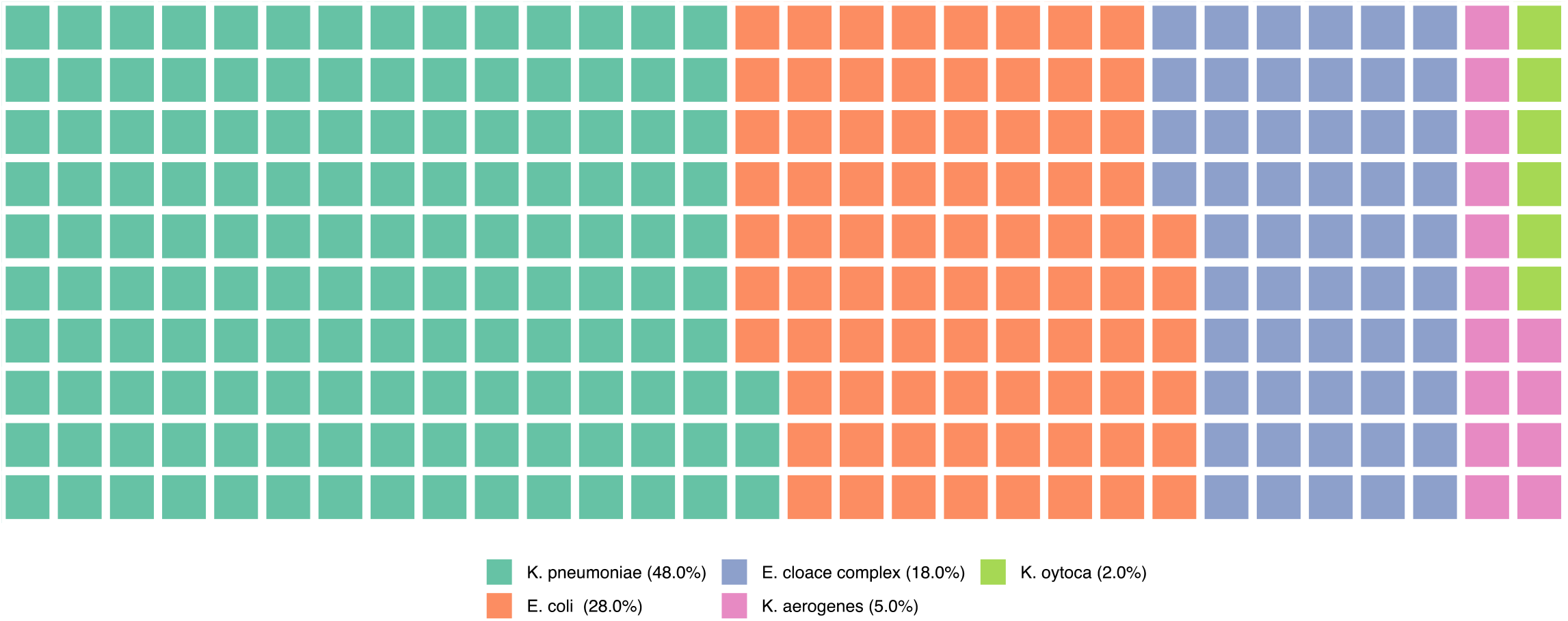
Waffle-plot of Carbapenem-resistant Enterobacterales cases by species collected by Georgia Emerging Infections Program between 2012-2017.

**Supplementary Figure 2.**
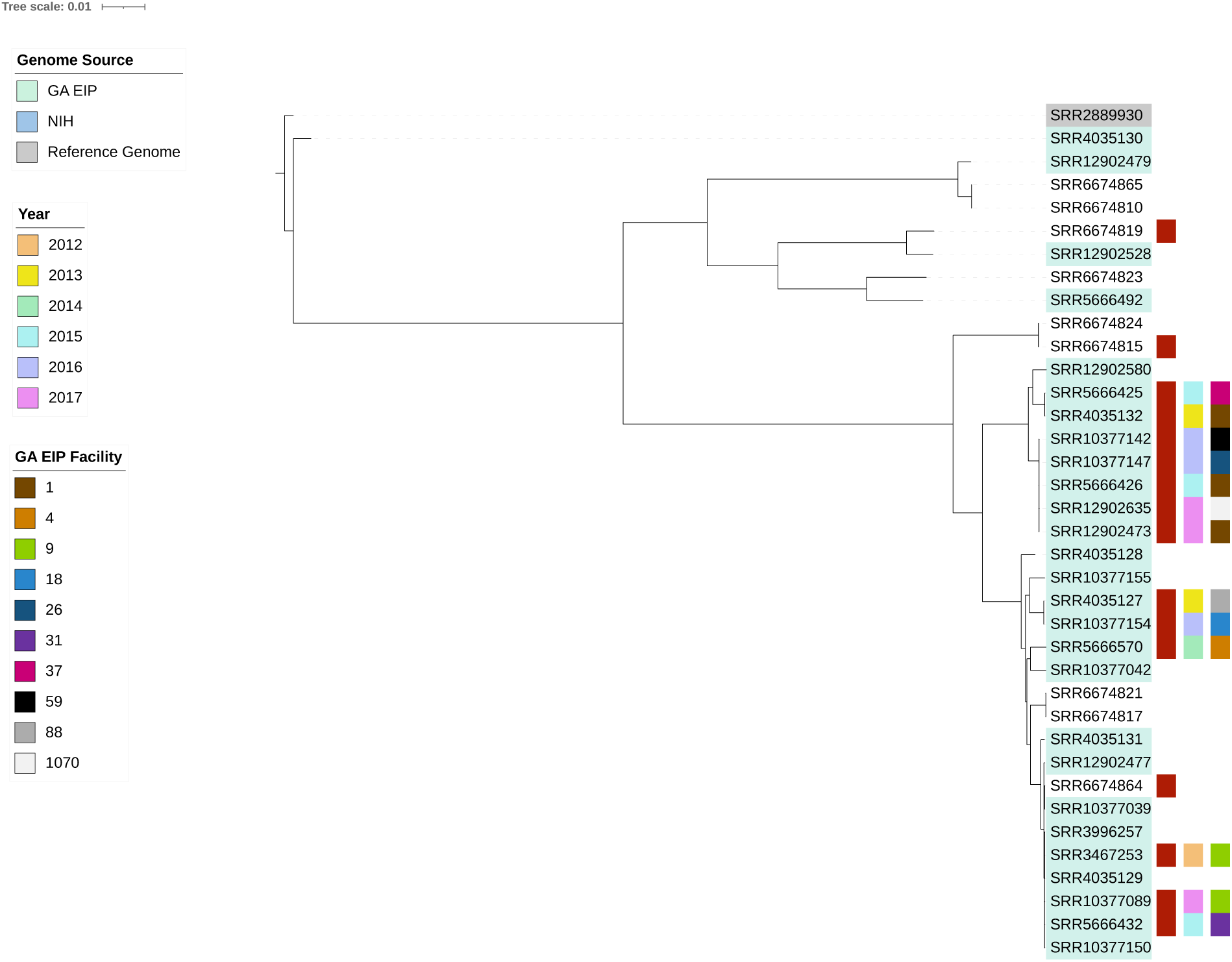
Phylogeny of *mcr-9* positive (n=13) and *mcr-9* negative (n=14) *E. cloacae* complex from Georgia Emerging Infections program (GA EIP) in addition to 9 available *E. cloacae* complex (three *mcr-9* positive, six *mcr-9* negative) from the National Institutes of Health. A phylogenetic tree based on a core gene alignment containing 1,904 genes defined using Roary v3.13.0. was generated using IQtree v2.0.3. A maximum likelihood tree was generated by running 1,000 bootstrap replicates under the generalized time-reversible model of evolution. The tree was visualized and Facility IDs and year of isolation were annotated using Interactive Tree of Life (iTOL) v4. for GA EIP *mcr-9* positive genomes only. Abbreviations: GA EIP: Georgia Emerging Infections Program, NIH: National Institutes of Health

**Supplementary Figure 3.**
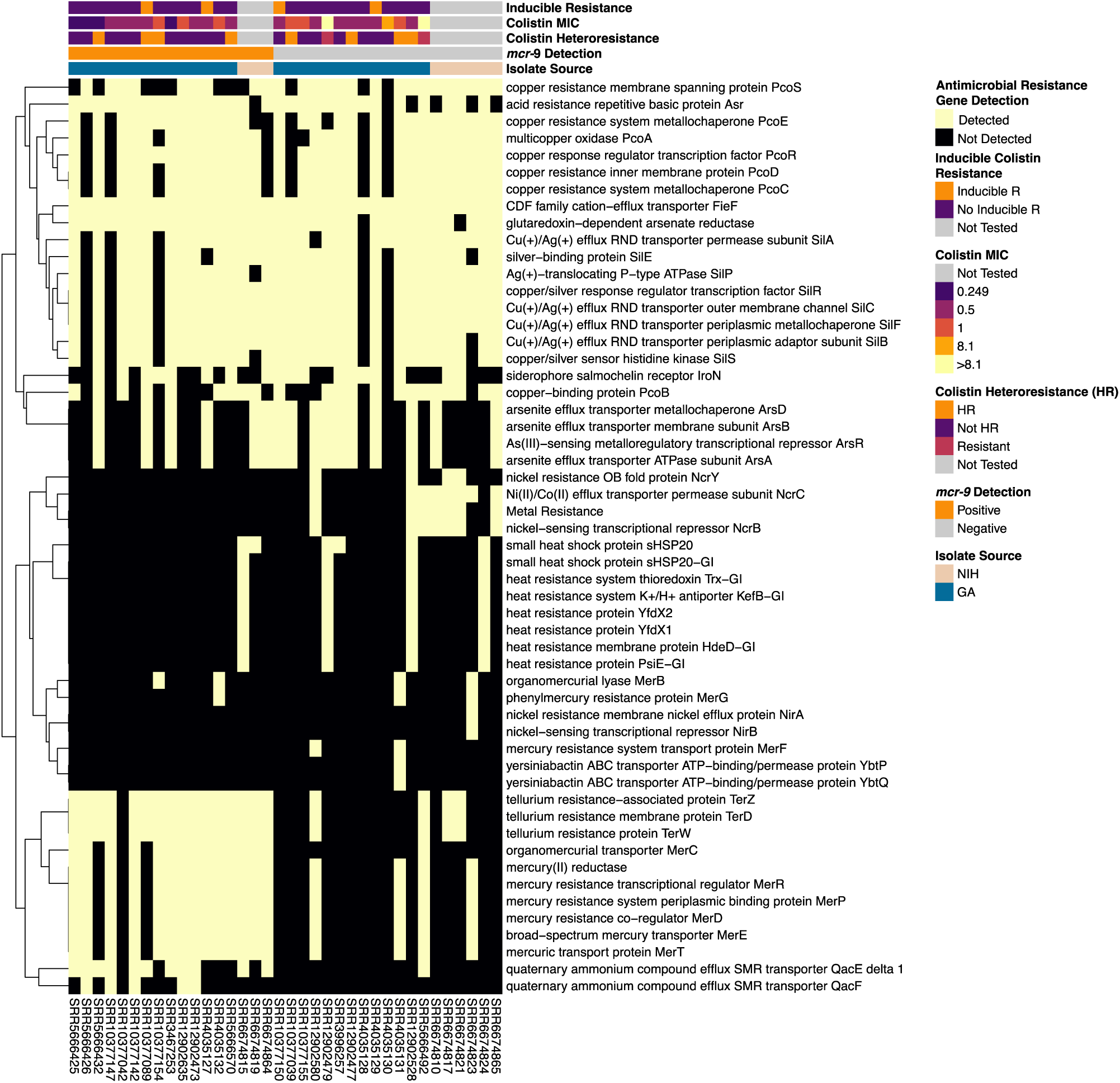
Genomes were annotated using Prodigal and virulence gene content was assessed using AMRFinder. Virulence gene presence/absence heatmaps were created using the package pheatmap on R version 4.0.2 (Vienna, Austria) and the RStudio interface version 1.3.1073 (Boston, MA, USA).

